# Phase 1 Study of Safety, Reactogenicity, and Immunogenicity of a Novel MVA-SARS-2-ST Vaccine Candidate Administered as Inhalation Boost in SARS-CoV-2 Immunized Adults

**DOI:** 10.1101/2025.10.07.25337492

**Authors:** Jens M. Hohlfeld, Swantje Hammerschmidt, Thomas Kayser, Anna Kutschenko, Anika Buchholz, Marc Permanyer, Lennart Riemann, Christoph Schindler, Antonia Zapf, Mahnaz Badpa, Verena Krähling, Stephan Becker, Christine Falk, Simon Schröder, Gerd Sutter, Asisa Volz, Reinhold Förster

## Abstract

**Background:** Existing parenteral SARS-CoV-2 vaccines protect against severe disease but do not reliably prevent infection or reinfection. Inhaled vaccines may elicit localized immunity in the respiratory tract, a principal entry site for SARS-CoV-2.

**Methods:** In this investigator-initiated, single-center, open-label phase 1 trial (NCT05226390, ClinicalTrials.gov), 23 healthy adults previously immunized with EU-approved SARS-CoV-2 vaccines received a single inhaled dose of Modified Vaccinia virus Ankara-(MVA)-SARS-2-ST (1×10^7^ IU), engineered to express a prefusion-stabilized SARS-CoV-2 spike. Participants were followed for 140 days. Primary endpoints included solicited local and systemic reactogenicity through day 7, unsolicited adverse events through day 28, serious adverse events throughout, and changes in spirometry and laboratory parameters. Secondary endpoints were changes in SARS-CoV-2 S1-specific IgG in serum and bronchoalveolar lavage; exploratory endpoints included changes in S1-specific IgA in serum and bronchoalveolar lavage, and methacholine responsiveness.

**Results:** No serious adverse events were observed. Over 28 days, mild or moderate adverse events occurred in 87% of participants, predominantly cough, headache, and fatigue, all resolved. Pulmonary function and methacholine responsiveness were stable, except for one transient 20% decrease in FEV1 on Day 14 that normalized subsequently. Serum IgG responses remained minimal, whereas a subset displayed increased bronchoalveolar IgA.

**Conclusion:** A single inhaled booster dose of MVA-SARS-2-ST was safe and generally well tolerated. While systemic antibody levels did not rise substantially, the observed mucosal IgA response in some participants points to a localized mucosal effect. Further studies are warranted to clarify underlying mechanisms and the significance of this response in diverse populations.

## Introduction

The identification of SARS-CoV-2 in Wuhan, China, in late 2019 marked the beginning of a global pandemic that led to millions of infections and deaths.^1^ In response, vaccines using viral vector platforms and mRNA technology were quickly developed and administered intramuscularly, which helped reduce mortality rates.^2^ However, these vaccines did not provide long-term protection against infection or reinfection, highlighting the need for improved vaccine solutions.^2^ Given the respiratory tract’s role as the primary gateway for viral entry, inhaled vaccines have emerged as a promising strategy to enhance mucosal immunity and stimulate bronchus-associated lymphoid tissue, potentially providing superior protection against respiratory pathogens like SARS-CoV-2.^3,4^

Modified Vaccinia virus Ankara (MVA) is a replication-deficient poxviral vector extensively evaluated in clinical settings, including thousands of recipients of MVA-Bavarian-Nordic, a licensed smallpox and Mpox vaccine. Furthermore, as demonstrated by tuberculosis vaccine trials showing that inhaled MVA was both well tolerated and effective, the lungs represent a compelling immunization route for MVA-based formulations.^5^ For SARS-CoV-2, two closely related MVA-based vaccines were developed: MVA-SARS-2-S (native spike sequence) and MVA-SARS-2-ST (prefusion-stabilized spike with cleavage site inactivation). While both underwent successful phase 1 clinical testing and were deemed safe, MVA-SARS-2-ST elicited stronger immune responses, though still lower than those induced by licensed mRNA or adenoviral vector vaccines.^6,7^ Preclinical studies further indicated that MVA-SARS-2-ST, used in an intramuscular priming and intranasal boosting regimen, offers complete protection against SARS-CoV-2 in animal models.^8^

Based on these findings, the present clinical trial assessed MVA-SARS-2-ST given by inhalation as a booster for adults primed with EU marketed SARS-CoV-2 vaccines, evaluating safety, reactogenicity, and immunogenicity.

## Methods

### Study Design and Participants

This investigator-initiated, single-center, open-label, Phase 1 trial was conducted at Hannover Medical School, Hannover, Germany, from June 2022 through November 2023. The study was registered prospectively with ClinicalTrials.gov (NCT05226390). Eligible participants were healthy adults aged 18 to 60 years who had previously received primary immunization with two doses of any EU marketed SARS-CoV-2 vaccine (mRNA-, vector-, protein-based, or attenuated virus) or a single dose of COVID-19 Vaccine Janssen, followed by a booster with any EU marketed mRNA vaccine. At screening, participants were required to have a SARS-CoV-2-specific IgG concentration of 10 to 1200 RU/mL by Anti-SARS-CoV-2-QuantiVac-ELISA (IgG). Full criteria are detailed in Supplementary Information S1. Written informed consent was mandatory for all participants, and the trial procedures adhered to Good Clinical Practice guidelines, applicable regulations, and sponsor operating procedures. The protocol (EudraCT no. 2020-004010-35) was reviewed and approved by the independent Ethics Committee of Hannover Medical School (no. 10012_AMG_mono_2021), Hannover, Germany and Paul-Ehrlich-Institute (4668/01), Langen, Germany.

### Procedures

Following screening (Day -28 to -1) and baseline assessments (Day -7 to -2), a single booster dose of 1×10^7^ ± 0.5 log MVA-SARS-2-ST in 0.5 mL was administered via inhalation on Day 0 (Figure 1). Over the 140-day follow-up, participants were monitored with spirometry, laboratory testing, vital signs, and adverse event assessment. Methacholine challenge testing was conducted before vaccination and again on Days 7 and 140. Pharmacodynamic analyses included repeated blood sampling, and bronchoscopy at baseline and Day 14 for collection of bronchoalveolar lavage (BAL) specimens.

**Figure 1:**
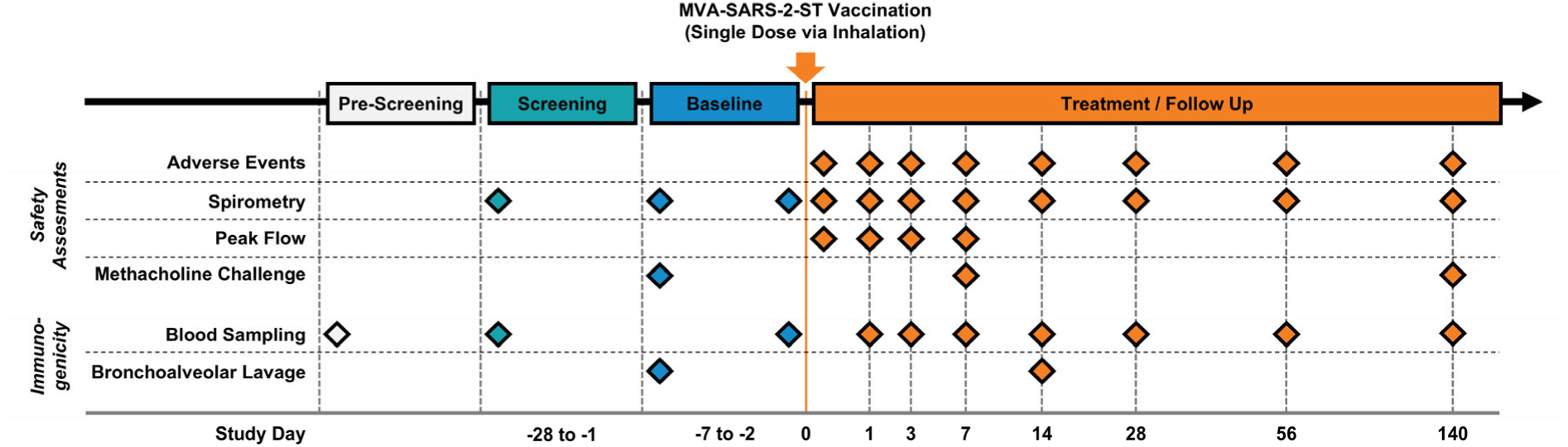
Overview of Study Procedures and Timeline. Participants underwent pre-screening, screening, and baseline evaluations before receiving a single inhaled booster dose of MVA-SARS-2-ST on Day 0. Safety assessments included monitoring for treatment-emergent adverse events, spirometry, peak flow measurement, and methacholine challenge testing. Immunogenicity analyses included measurement of SARS-CoV-2 S1-specific IgG and IgA antibodies in serum and bronchoalveolar lavage (BAL). Assessments were performed at baseline and at specified time points during the 140-day follow-up, as depicted in the figure.

### Inhalation Method and Vaccine Aerosol Stability

To generate the vaccine aerosol, 0.5 mL of MVA-SARS-2-ST was loaded into a vibrating mesh nebulizer (M-neb dose+, NEBU-TEC, Elsenfeld, Germany). Participants wore a nose clip and performed approximately 70-90 breath-actuated inhalations, each delivering ∼7 µL of aerosol, followed by a 5-second breath hold and exhalation through a dedicated port.

To confirm that this nebulization method preserved vaccine viability, we performed in-use stability assessments. Given the need for rapid data during the evolving pandemic, these tests were completed using MVA-SARS-2-S instead of MVA-SARS-2-ST; both constructs share the same poxviral backbone. Nebulized and non-nebulized samples were compared by measuring infectious titers in DF-1 cells, as detailed in Supplementary Information S2. Weight differences before and after nebulization were used to quantify the amount of vaccine delivered, retained in the nebulizer, or lost.

### Adverse Events

All adverse events (AEs) were systematically recorded throughout the study. Solicited local and systemic adverse events, collected during the first 7 days post-vaccination, were specifically assessed to evaluate reactogenicity. In addition, adverse events of special interest (AESIs) were monitored for the duration of the trial and included generalized convulsions, acute disseminated encephalomyelitis, Guillain-Barré syndrome, thrombocytopenia, anaphylaxis, and vasculitides.

### Pulmonary Function Testing

Spirometry and methacholine challenge followed standardized guidelines.^9,10^ Outcomes included forced expiratory volume in 1 second (FEV1), forced vital capacity (FVC), and the methacholine concentration provoking a 20% decrease in FEV1 (PC20).

### Bronchoscopy

Bronchoscopic procedures adhered to published methods.^11^ Participants received a bronchodilator (200 µg salbutamol inhalation), sedation (midazolam 0.05-0.1 mg/kg IV), and topical anesthesia (lidocaine). BAL was obtained from the right middle lobe or lingula by instilling four 50-mL aliquots of pre-warmed sterile 0.9% saline.

### Immunologic Analysis

BAL and blood samples were processed according to standard laboratory protocols. We measured SARS-CoV-2 specific IgG for the screening with the commercial Anti-SARS-CoV-2 QuantiVac ELISA (IgG) (Euroimmun, EI 2606-9601-10G) and for the longitudinal study analysis with an in-house ELISA^12^ approved by Paul-Ehrlich-Institute. IgA was analyzed using a commercial bead-based assay (Millipore, HC19SERA1-85K-04). For the bead-based assay, plasma was diluted 1:250 and BAL 1:5. The semi-quantitative readout is given as median fluorescence intensity (MFI) of > 50 beads for each antigen and sample, acquired by the Bio-Plex 200 machine and the Bio-Plex Manager™ Version 6.0 software (Bio-Rad, Hercules, USA).

### Endpoints

Primary endpoints included safety and reactogenicity, measured as solicited local and systemic symptoms through day 7, unsolicited adverse events through day 28, serious adverse events at any time, and changes in spirometry (FVC, FEV1, FEV1/FVC, peak flow) or laboratory values at predefined intervals. Secondary endpoints involved changes from baseline in SARS-CoV-2 S1-specific IgG in serum (days 7, 14, 28, 56, 140) and BAL (day 14). Exploratory endpoints included SARS-CoV-2 S1-specific IgA in serum (days 7, 14, 28, 56, 140) and BAL (day 14), and methacholine challenge (PC20) at days 7 and 140.

### Statistical Analysis and Sample Size

No formal sample size calculation was performed, as is typical for Phase 1 studies. A cohort size of 24 participants was deemed sufficient to allow descriptive evaluation of safety and immunogenicity, and to estimate a two-sided 95% confidence interval with a precision of 0.42 for an event rate of 50% or at least 0.24 for rare events (i.e., one event in 15 participants), as calculated using Confidence Intervals for One Proportion (PASS 15). The evaluated for safety (EFS) set included all vaccinated participants, whereas the per-protocol (PP) set excluded individuals with major protocol deviations. All analyses were of descriptive nature. It was not intended to test hypotheses in a confirmatory sense. Data were summarized by number of non-missing values, means with two-sided 95% confidence interval, medians, standard deviations, coefficients of variance, minimum, maximum and interquartile ranges or absolute and relative frequencies, as appropriate. Missing values have not been imputed.

## Results

### Vaccine Candidate Integrity After Aerosolization

Assessment of vaccine integrity following nebulization demonstrated a reduction in infectious titer from 2.77×10^7^ IU/mL to 1.85×10^7^ IU/mL, corresponding to a 0.17 log reduction (Figure 2A). Volumetric analysis revealed that 87% of the vaccine was recovered in the collection vessel, ∼8% remained in the nebulizer, and ∼5% was irrecoverably lost (Figure 2B). Additionally, MVA-SARS-2-S titers within the nebulization device declined over time at room temperature, culminating in a 0.165 log-titer reduction after two hours (mean half-life = 3.51 hours; Figure 2C). Overall, the cumulative loss of activity from both titer drop, and volume loss was 42%, which corresponds to an approximate 0.24 log reduction. Nevertheless, the final product titer was routinely sufficient to ensure delivery of 1.00×10^7^ IU per dose (Supplementary Table S1).

**Figure 2:**
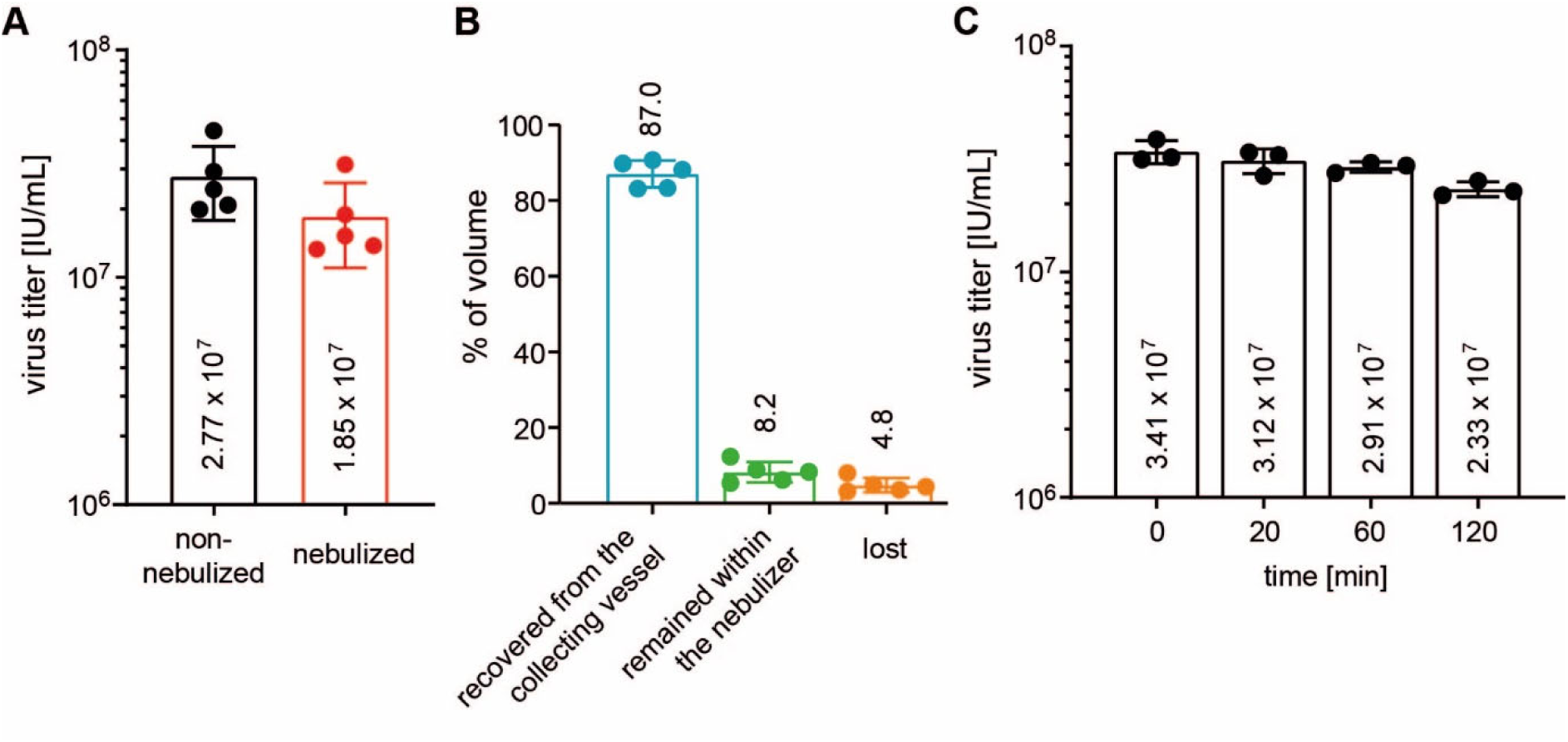
In-use Stability of MVA-SARS-2-S. (A) Effect of nebulization on MVA-SARS-2-S titers (non-nebulized vs. nebulized). (B) Volumetric recovery after nebulization, indicating proportions recovered from the collecting vessel, retained in the nebulizer, or lost. Panels A and B present mean ± SD from five virus aliquots tested in three independent experiments; numerals within bars show mean values or mean percentages. (C) Stability over time at room temperature, with titers measured at the indicated time points. Panel C presents mean ± SD from three virus aliquots tested in three independent experiments.

### Trial Population

A total of 124 participants were screened for SARS-CoV-2-specific IgG in a pre-screening phase; 41 met the antibody concentration requirements of 10 to 1200 RU/mL. Eighteen of these 41 individuals were excluded for failing to meet other inclusion criteria, leaving 23 participants who received the inhaled booster (see Figure 3). All 23 completed the study and were included in both the safety (EFS) and per-protocol (PP) analyses. Participant demographics are summarized in Table 1, showing a predominantly female (78.3%) and White (100%) cohort, with a mean age of 35.3 years and a mean BMI of 25.5 kg/m².

**Figure 3:**
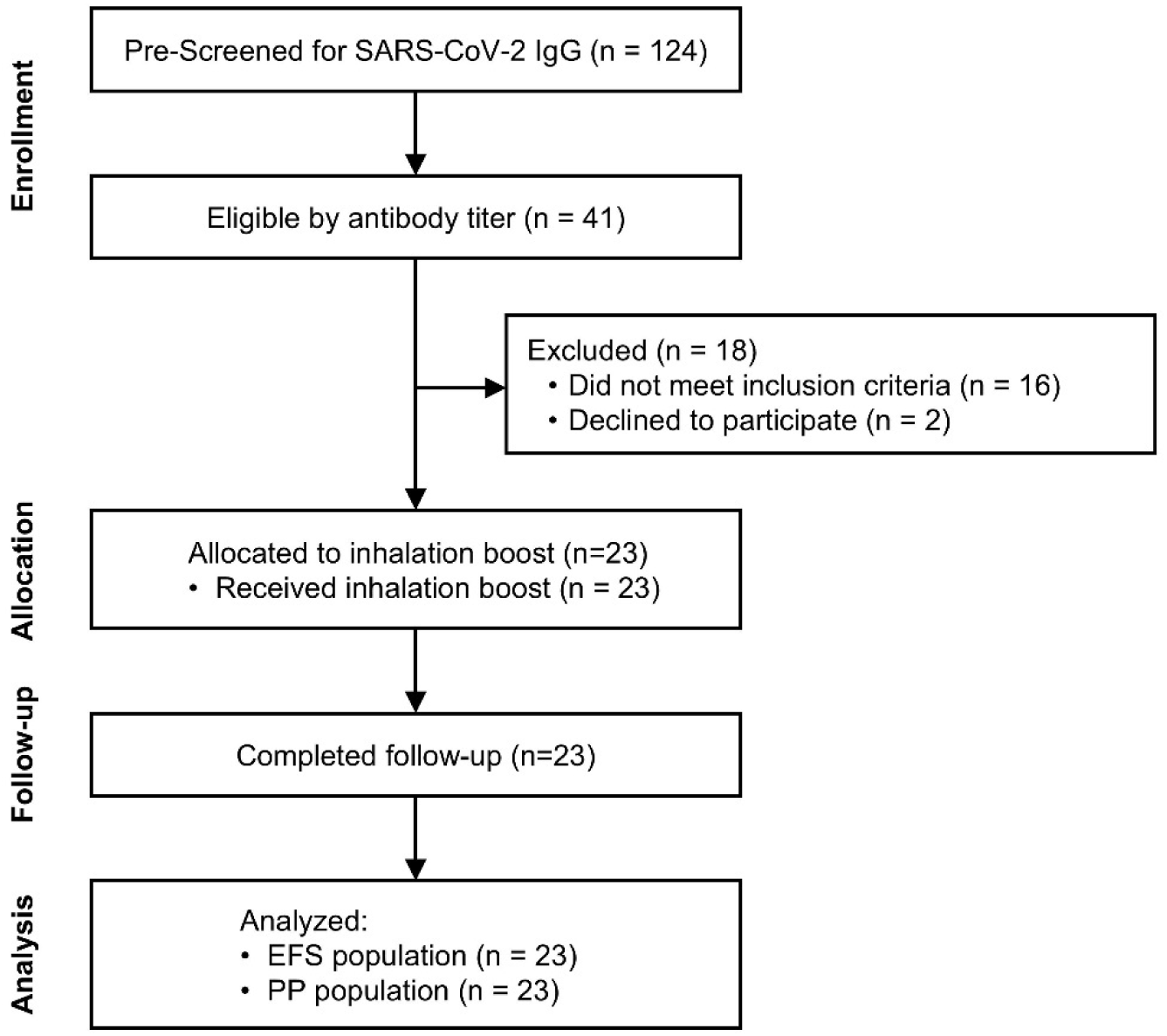
Participant Disposition Throughout the Study. A total of 124 participants were pre-screened, with 41 meeting the antibody titer criterion. Following exclusion, 23 participants received the inhaled MVA-SARS-2-ST booster, completed the study, and were included in both safety (EFS) and per-protocol (PP) analyses.

**Table 1.** Demographics and Baseline Characteristics of Enrolled Participants (EFS, N=23). Data are presented as mean ± SD [range] for continuous variables and n (%) for categorical variables. BMI indicates body mass index; RU/mL, relative units per milliliter.

| Variable | Summary Statistics |
| --- | --- |
| Age, years | 35.3 ± 11.0 [21-57] |
| Sex, n (%) |  |
| Female | 18 (78.3) |
| Male | 5 (21.7) |
| Height, cm | 172.3 ± 11.3 [147-196] |
| Weight, kg | 75.8 ± 12.6 [56-108] |
| BMI, kg/m<sup>2</sup> | 25.5 ± 2.9 [20-29] |
| Anti-SARS-CoV-2 S1-specific IgG, RU/ml | 390.9 ± 294.3 [45-1171] |

### Reactogenicity

Solicited local and systemic reactogenicity were recorded for 7 days post-vaccination and are summarized in Figure 4. Cough was the most common local adverse event (occurring in fewer than 25% of participants), whereas chest discomfort, chest pain, and dry throat each affected fewer than 5% (1 out of 23 participants). Throat irritation occurred in fewer than 10% (2 out of 23 of participants). All local events were mild to moderate (Grade 1 or 2). Systemic events, including abdominal pain, hyperhidrosis, malaise, and myalgia, were each reported by fewer than 10% of participants, while pyrexia, headache, or fatigue ranged from 17.4 to 26.1%. Again, these systemic events were of mild or moderate intensity, and no Grade 3, 4, or 5 reactogenicity was observed.

**Figure 4.**
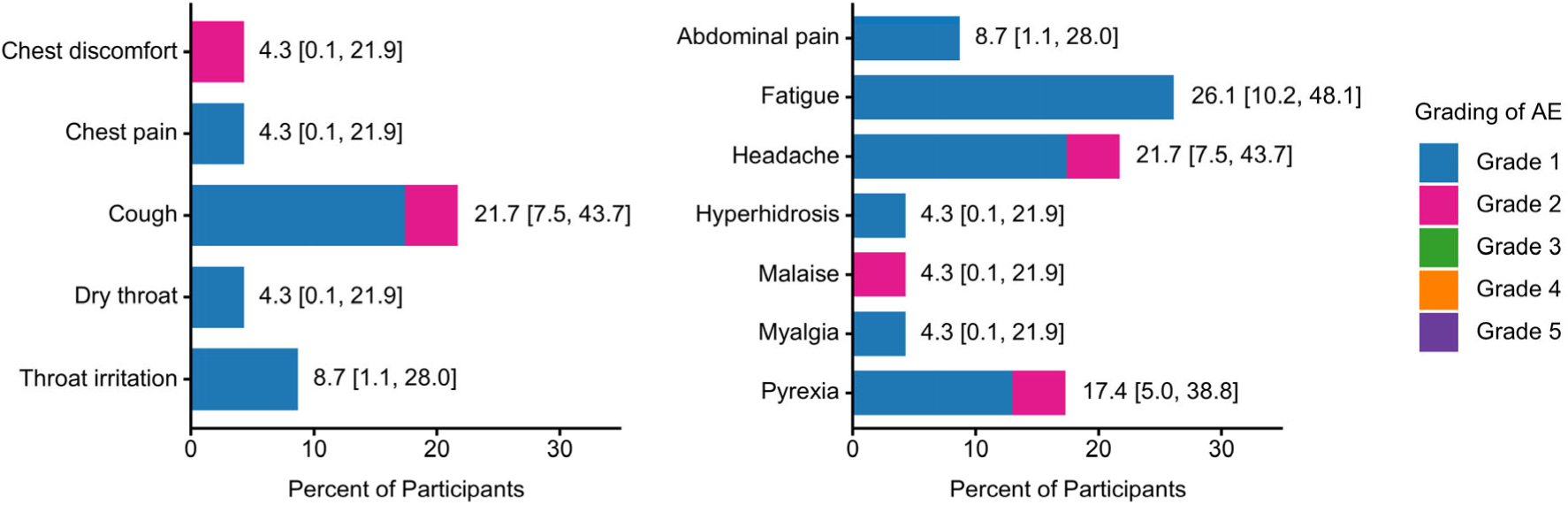
Solicited Local (Left Panel) and Systemic (Right Panel) Reactogenicity. Bars represent the proportion of participants reporting each MedDRA preferred term within 7 days, categorized by severity (highest grade counted if multiple). Two-sided 95% confidence intervals indicate the total incidence across all severity grades (Grade 1 = Mild, Grade 2 = Moderate, Grade 3 = Severe, Grade 4 = Life-threatening, Grade 5 = Death).

### Safety

During the primary 28-day observation period, 87% of participants reported at least one unsolicited adverse event (AE), totaling 65 events. The most common AEs within this period were cough (26.1% of participants), headache (21.7%), fatigue (17.4%), and hypertension (17.4%) (Table 2). With the exception of one participant who experienced a single-day episode of severe epistaxis (Grade 3, unlikely related to treatment), all events were of mild or moderate intensity (Grade 1 or 2). No serious adverse events and no clinically significant laboratory abnormalities were observed.

**Table 2.**
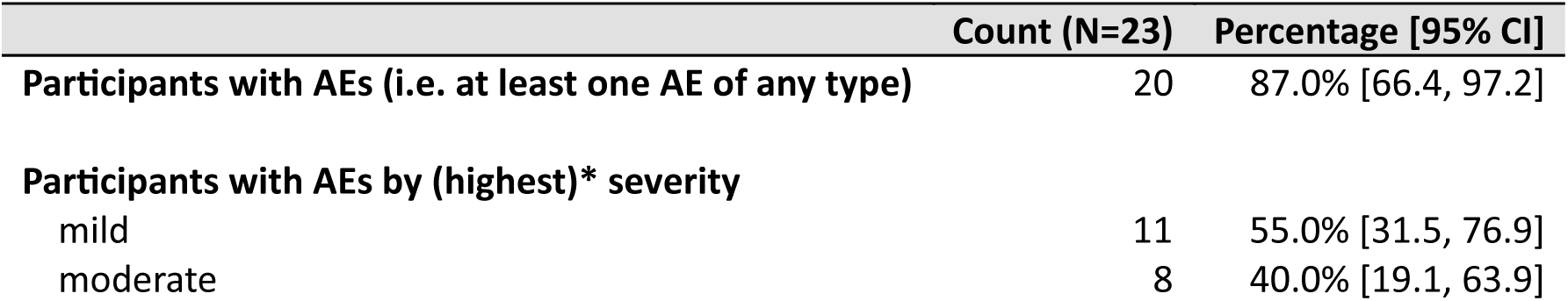
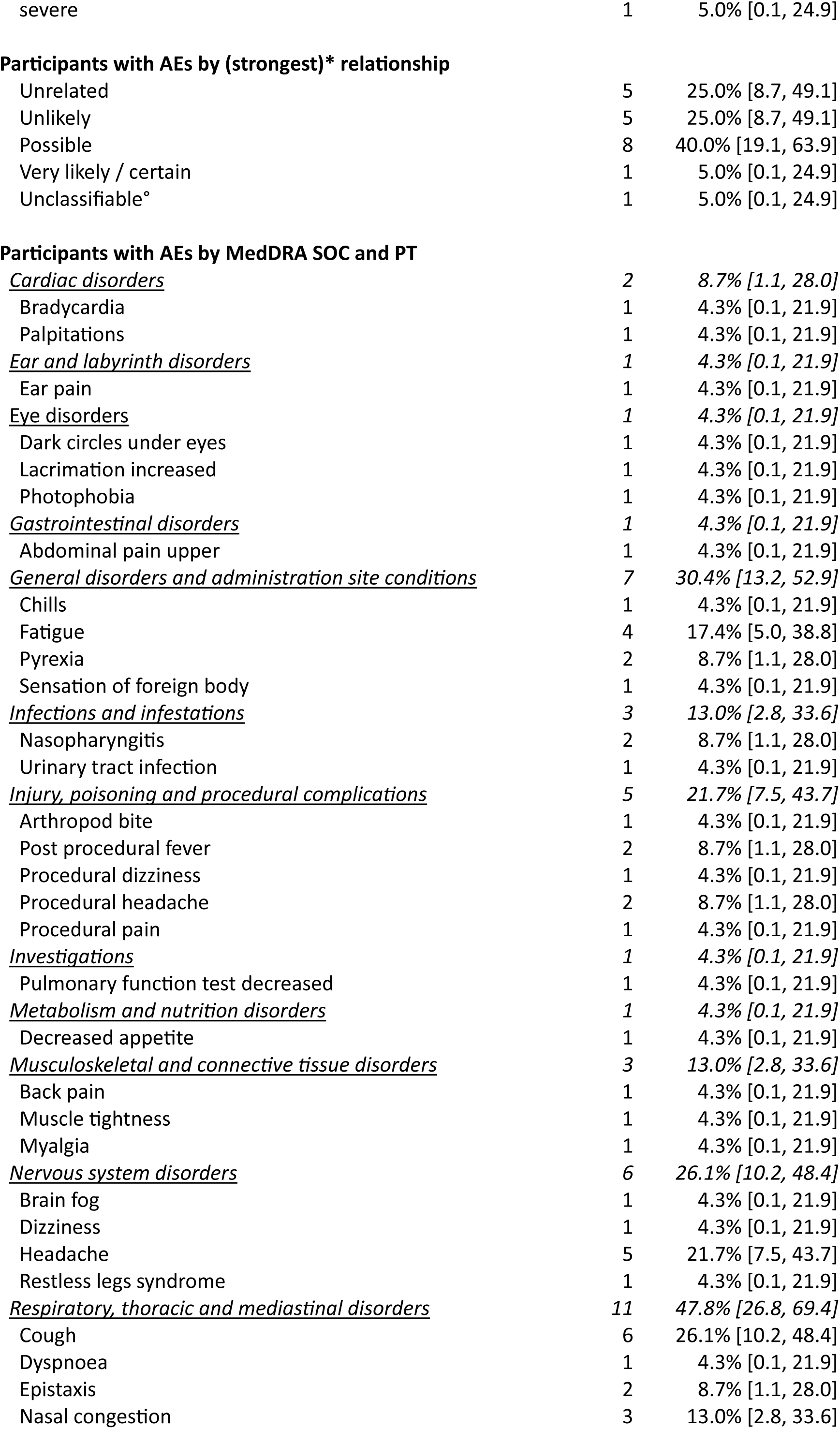
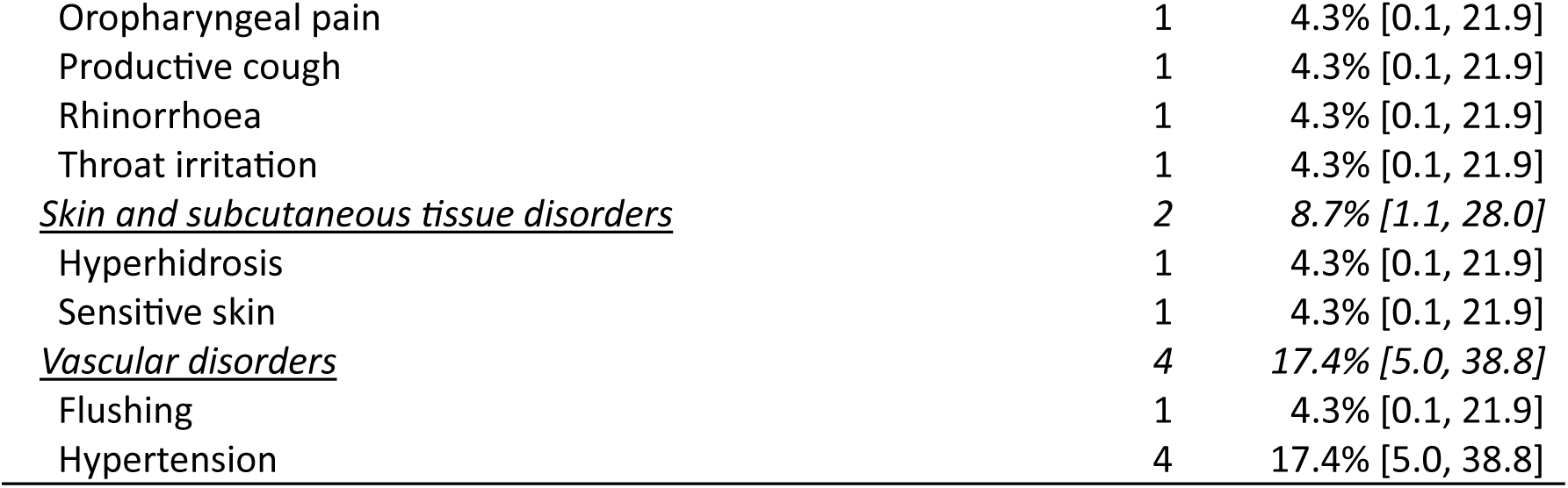
Unsolicited Adverse Events Within 28 Days After Inhaled MVA-SARS-2-ST Vaccination. Adverse events are tabulated by MedDRA system organ class (SOC) and preferred term (PT), showing the number and percentage of participants (EFS, n = 23) with at least one event of each type. Absolute (Count) and relative (Percent) frequencies, as well as 95% confidence intervals (CI) are reported. * For participants with multiple adverse events, only the highest severity or the strongest relationship is counted. ° The strongest classifiable relationship for this participant was ‘Unlikely’.

Over the entire study period, 91.3% of participants reported a total of 147 AEs. A comprehensive listing of all reported AEs throughout the study, sorted by participant, is provided in Supplementary Table S2.

### Pulmonary Function Outcomes

Serial spirometry measurements revealed no clinically concerning changes in FEV1 or FVC. The greatest mean percentage decrease in either parameter was 2.14% at Day 14 (Figure 5A and B), corresponding to mean absolute changes of approximately 0.093 L for FEV1 and 0.110 L for FVC. One participant had a clinically notable FEV1 drop of −0.84 L (−20.19%) on Day 14, which resolved to baseline by study conclusion (Figure 5C and D).

**Figure 5.**
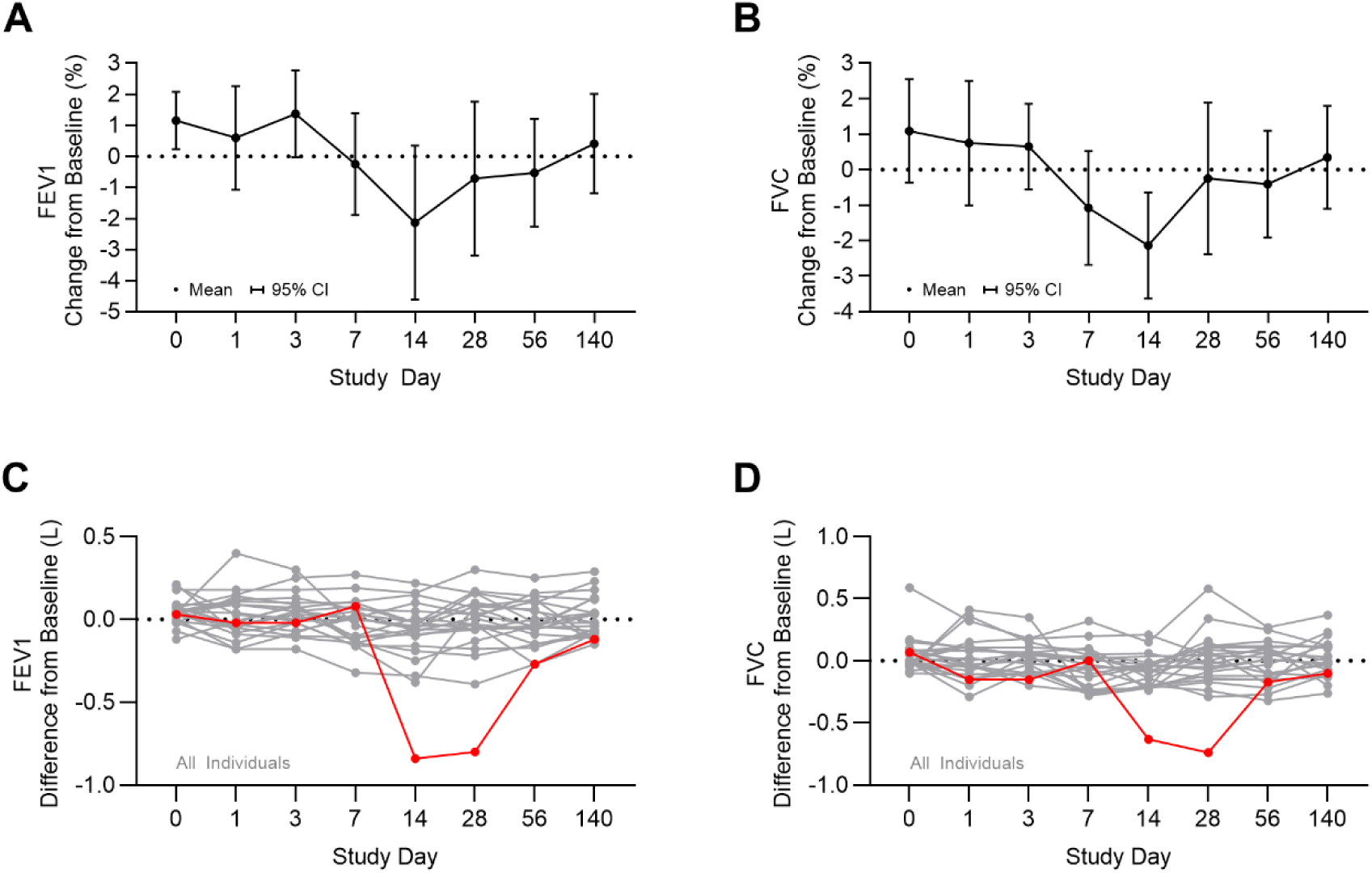
Change from Baseline in Pulmonary Function After Inhaled MVA-SARS-2-ST Vaccination. (A, B) Percent change from baseline in forced expiratory volume in 1 second (FEV1, A) and forced vital capacity (FVC, B) shown as mean with 95% confidence intervals at each time point. (C, D) Change from baseline in FEV1 (C) and FVC (D) in liters for individual participants; with one participant experiencing a transient decrease at Day 14 (red line). Day 0 is post-vaccination, Day 14 is pre-bronchoscopy.

Methacholine challenge testing yielded normal mean PC20 values at baseline (31.0 ± 4.8 mg/mL), Day 7 (30.2 ± 6.1 mg/mL), and Day 140 (32.0 ± 0 mg/mL), confirming stable bronchial responsiveness to methacholine.

Overall, these observations confirm the absence of clinically meaningful respiratory changes, with no evidence of obstruction or lasting functional deficits.

### Immunogenicity

Compartment-specific analysis of humoral immunogenicity following inhaled vaccination demonstrated modest and variable changes across the cohort (Figure 6).

**Figure 6.**
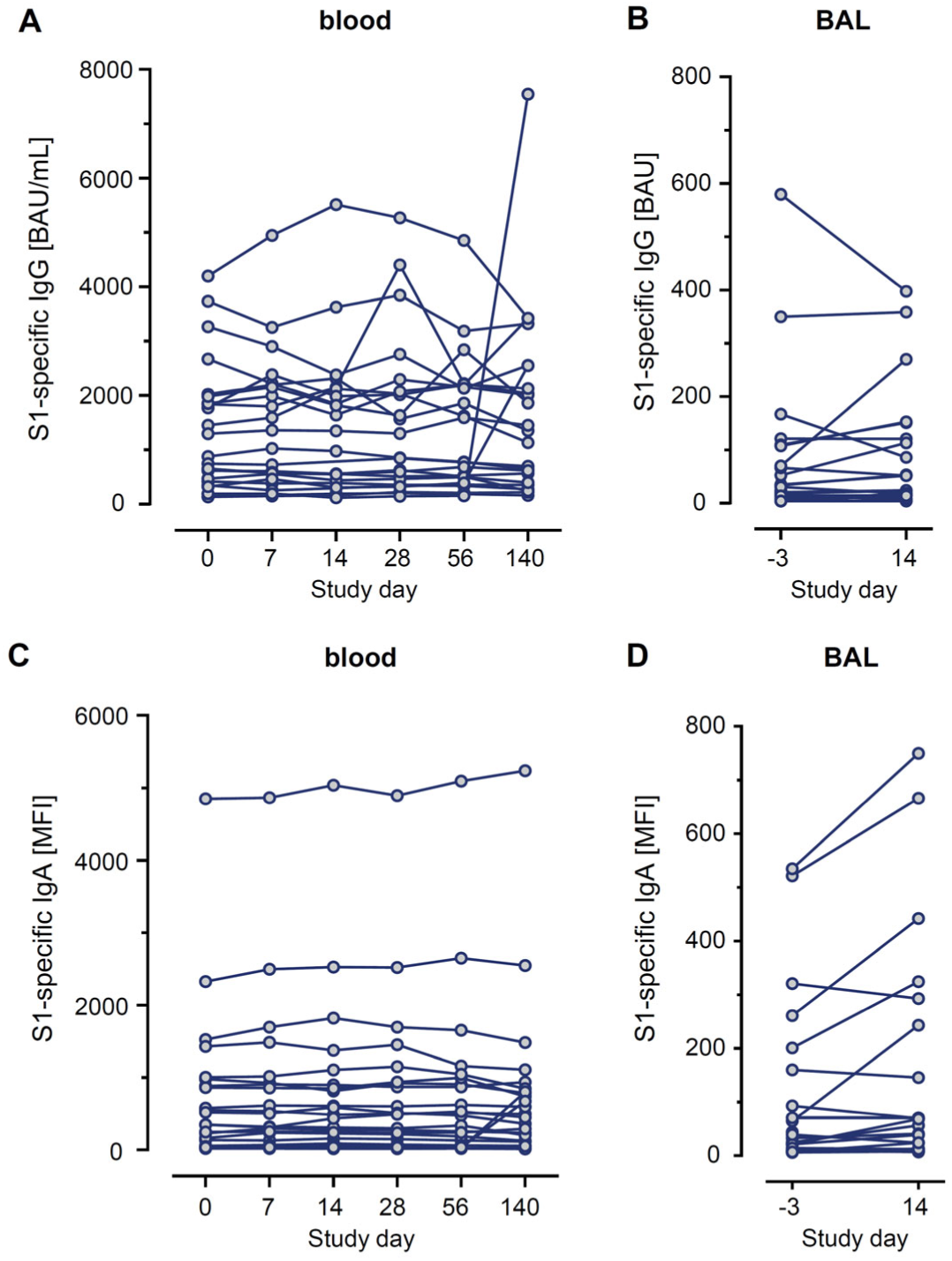
Levels of IgG (upper panels) and IgA (lower panels) binding antibodies to SARS-CoV-2 spike S1 protein in serum (left panels) and bronchoalveolar lavage (BAL, right panels) at designated study days following inhaled vaccination. Each line represents an individual participant. Summary statistics are provided in Supplementary Tables S3 through S6.

In serum, S1-specific IgG showed positive mean changes at Day 28 and Day 140, with median changes near zero or negative (Day 28 mean change, 130.8 BAU/mL; median change, −1.0 BAU/mL; Day 140 mean change, 316.7 BAU/mL; median change, −62.0 BAU/mL; Supplementary Table S3; Figure 6A). Of note, the positive mean change at Day 140 coincided with four participants identified as having suspected breakthrough SARS-CoV-2 infection after Day 56, based on the combined findings of reported COVID-19 as an adverse event, a positive PCR test for SARS-CoV-2, or concurrent rises in NCP-specific IgG (Supplementary Table S7 and Supplementary Figure S1). However, the Day 28 mean increase preceded these events and likely reflects individual variability. Serum S1-specific IgA exhibited only minor changes overall, with a transient rise at Day 28 that was not sustained (Day 28 mean change, 33.5 MFI; Day 28 median change, 20.0 MFI; Supplementary Table S5; Figure 6C).

In BAL, S1-specific IgG remained unchanged at Day 14 (mean change, 6.0 BAU, 95% CI, −22.8 to 34.7; Supplementary Table S4; Figure 6B). By contrast, S1-specific IgA in BAL exhibited a moderate mean increase at Day 14 (mean change, 40.5 MFI; median change, 5.3 MFI; range, −128 to 215 MFI; Supplementary Table S6; Figure 6D), driven by a subset of participants with pronounced rises, while most showed small or no change.

Overall, the data indicate a compartmentalized mucosal IgA induction in some participants, with limited systemic antibody changes at the cohort level.

## Discussion

In this investigator-initiated, open-label, phase 1 study, the MVA-SARS-2-ST vaccine which was previously tested safe and efficacious as intramuscular injection^6,7^, was administrated via inhalation. The vaccine aerosol was generated using a vibrating-mesh nebulizer which allowed for functional preservation of virus activity and minimal technical losses as demonstrated by in-use stability assessment with an overall 0.24 log reduction of virus titer by nebulization. Inhaled MVA-SARS-2-ST vaccine demonstrated a favorable safety profile in healthy adults previously primed with standard SARS-CoV-2 vaccines. Only a small proportion of participants experienced solicited local or systemic reactogenicity, and these events were mild or moderate in intensity and self-limiting. Unsolicited AEs of any type were also mainly of mild and moderate intensity. Across the entire observation period, no serious adverse events or suspected unexpected serious adverse reactions were reported, and no participant experienced permanent harm.

Assessment of pulmonary function demonstrated a transient reduction in mean FEV1 and FVC at Day 14, with the magnitude of change closely approximating the minimal clinically important difference defined for these measures.^13^ However, this reduction was primarily attributable to a single participant who experienced a transient FEV1 decline exceeding 20%. Comprehensive clinical and laboratory evaluation of this individual revealed no signs of acute infection: C-reactive protein, leukocyte, and eosinophil counts remained within normal limits, and multiplex PCR testing for influenza A, influenza B, RSV, and SARS-CoV-2 was negative. The pulmonary function subsequently returned to baseline without sequelae. Importantly, methacholine challenge testing remained stable across all participants, a noteworthy finding given that immune responses to respiratory viruses have been associated with increased bronchial reactivity.^14^ Nevertheless, the pronounced individual event highlights the need for vigilant pulmonary monitoring and consideration of potential risks in individuals with preexisting respiratory conditions.

In terms of immunogenicity, our trial found that serum and bronchoalveolar IgG levels remained largely unchanged following inhaled vaccination, yet a distinct rise in BAL S1-specific IgA emerged in a subset of participants. Importantly, these mucosal IgA on Day 14 increases occurred before any reported or suspected breakthrough infections, which were recorded after Day 56, supporting the interpretation that these changes reflect a vaccine-associated effect rather than intercurrent infection. Although immunogenicity was not the primary endpoint, these data support the concept that inhaled vaccination may help trigger site-specific immune responses in the lower airways, albeit not uniformly across all participants. The mechanisms driving such variable IgA responses remain unclear, underscoring the need for subsequent, more detailed analyses. Of note, while the humoral immune responses reported here were part of the prespecified analysis, additional exploratory investigations of humoral and cellular immunity were conducted in parallel and will be reported separately to provide a comprehensive assessment of the immunologic effects of inhaled MVA-SARS-2-ST vaccination.

Several limitations should be acknowledged. First, being a first-in-human, phase 1 study, we enrolled a relatively small sample of participants with diverse pre-existing priming histories and varying baseline anti-spike protein titers (10-1200 RU/mL). Although small cohorts are commonly used to evaluate preliminary safety, the heterogeneity in preexisting immunity may have attenuated detectable immunogenic signals. Notably, prior data show that after a third standard vaccine dose, anti-spike levels on average reach 2000 RU/mL, later declining to an average of 348 RU/mL (146-594 RU/mL range).^15^ By enrolling individuals with ≤1200 RU/mL, we specifically targeted those who were in potential need of a booster and had room for immunologic improvement within the scope of this study.

Second, the absence of a concurrent placebo control could introduce bias in adverse event reporting, yet objective pulmonary parameters (spirometry, methacholine challenge) served to mitigate this limitation.

Third, this study was constrained by assessing only a single dosage strength. While there was convincing evidence from preclinical models that the tested dose and a tenfold higher dose induced a dose-dependent immune response that prevented animals from SARS-CoV-2 infections,^8^ the lack of formal toxicology assessments precluded further dose escalation in the present study.

In conclusion, this first-in-human trial demonstrates that a single 1×10^7^ IU booster dose of inhaled MVA-SARS-2-ST is safe and generally well tolerated in healthy adults with pre-existing antibody titers. Although systemic antibody levels did not show marked changes, a subset of participants demonstrated increased mucosal IgA in the lower respiratory tract. These findings suggest a potential for compartmentalized immunity at the site of viral entry, yet underscore the need for additional investigations to define the underlying mechanisms, optimize dosing strategies, and evaluate the persistence of these responses in larger and more diverse populations.

## Acknowledgements

The authors gratefully acknowledge the contributions of Thomas Hesterkamp from the Translational Project Management Office of the German Center for Infection Research, Braunschweig for clinical trial set-up activities, and the commitment of the clinical and technical staff involved in this study. The authors thank all volunteers for study participation and IDT Biologika, Dessau for investigational medicinal product supply.

## Data availability

The data that support the findings of this study are available from the corresponding author, JMH, upon reasonable request.

## Funding

This work was funded by the State of Lower Saxony Ministry of Science and Culture (MWK Niedersachsen) (14-76103-184 CORONA-11/20), by the German Center for Infection Research (Deutsches Zentrum für Infektionsforschung, DZIF) TTU 01.934 (grant no 8001801934) and TTU 01.941 (grant no 8001801941), by the German Research Foundation (Deutsche Forschungsgemeinschaft, DFG,) Excellence Strategy EXC 2155 “RESIST” (Project ID39087428), and by the German Center for Lung Research (Deutsches Lungenzentrum, DZL, grant 82DZL002B1).

## Author/Contributor Roles

CRediT, Contributor Roles Taxonomy

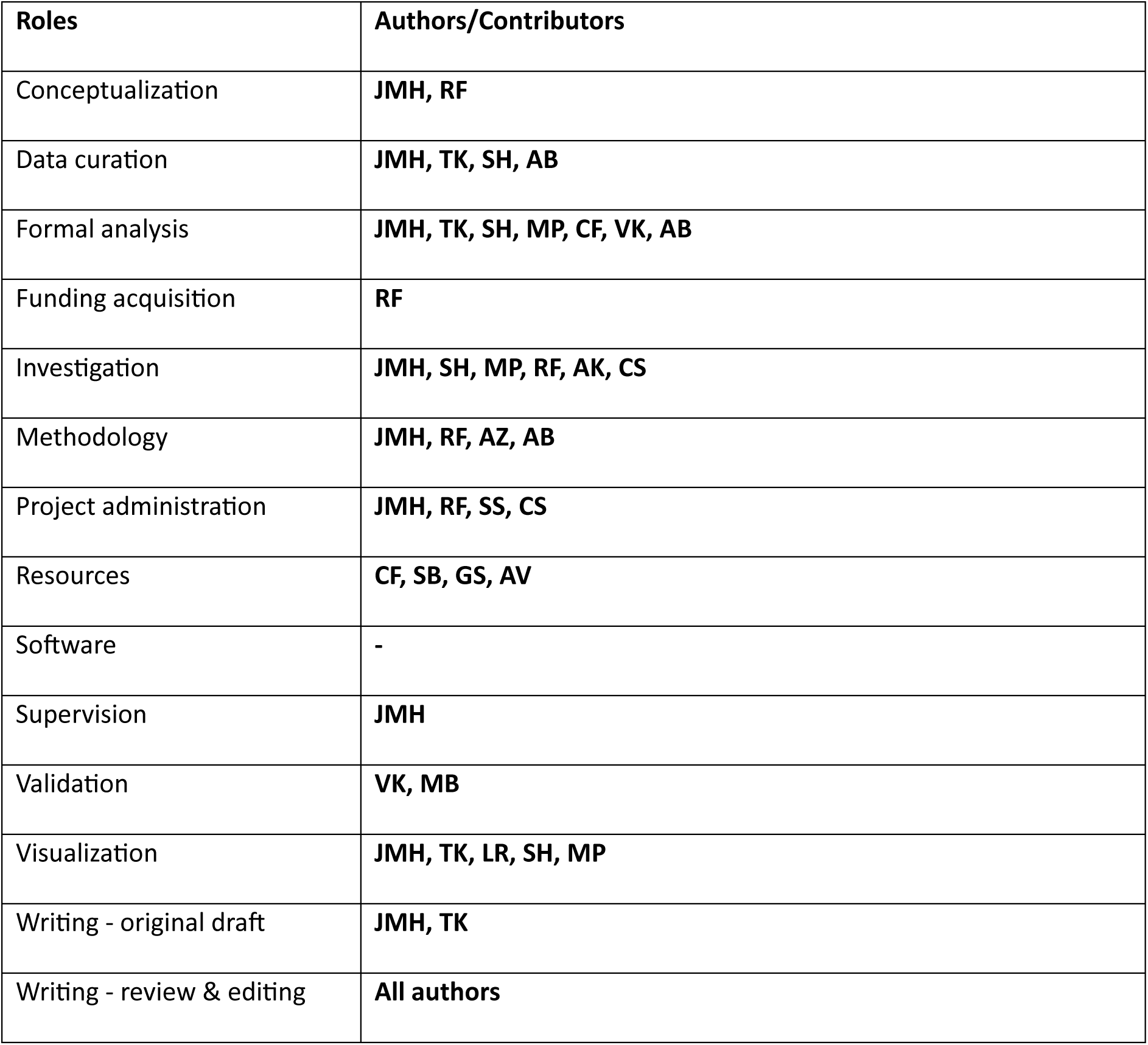

| **Roles** | **Authors/Contributors** |
| --- | --- |
| Conceptualization | JMH, RF |
| Data curation | JMH, TK, SH, AB |
| Formal analysis | JMH, TK, SH, MP, CF, VK, AB |
| Funding acquisition | RF |
| Investigation | JMH, SH, MP, RF, AK, CS |
| Methodology | JMH, RF, AZ, AB |
| Project administration | JMH, RF, SS, CS |
| Resources | CF, SB, GS, AV |
| Software | - |
| Supervision | JMH |
| Validation | VK, MB |
| Visualization | JMH, TK, LR, SH, MP |
| Writing - original draft | JMH, TK |
| Writing - review & editing | All authors |

## Conflict of Interest Statement

Jens M Hohlfeld reports grants for clinical trial conduct to his institution from Astellas Pharma GmbH, AstraZeneca, Bayer AG, Boehringer Ingelheim Pharma GmbH & Co. KG, Calibr at Scripps Research, Chiesi, CSL Behring, Desitin Arzneimittel GmbH, EpiEndo, F. Hoffmann-La Roche AG, Genentech, Inc., OM Pharma SA, ReAlta Life Sciences, Sanofi-Aventis Deutschland GmbH, and personal fees from Boehringer Ingelheim Pharma GmbH & Co. KG, Celerion, and Cureteq, all of which are outside the submitted work.

## Notes

### Clinical Trial

NCT05226390

### Author Declarations

Ethics Committee of Hannover Medical School gave ethical approval for this work (no. 10012_AMG_mono_2021).

