## Supplemental Material for "Phase 1 Study of Safety, Reactogenicity, and Immunogenicity of a Novel MVA-SARS-2-ST Vaccine Candidate Administered as Inhalation Boost in SARS-CoV-2 Immunized Adults"

#### Supplementary Information S1: Detailed Inclusion and Exclusion Criteria for Study Participants

Eligible participants were healthy male or female adults aged 18 to 60 years who had received primary immunization with either two doses of any EU marketed SARS-CoV-2 vaccine (mRNA, vector, protein-based, or attenuated virus) or a single dose of COVID-19 Vaccine Janssen, followed by a booster with any EU marketed mRNA vaccine. Additional requirements included anti-SARS-CoV-2 IgG levels between 10 and 1200 RU/mL, normal pulmonary function, and a BMI of 18.5 to 30.0 kg/m². Individuals with prior MVA vaccination, pulmonary disease, immunodeficiency, or other significant medical conditions were excluded.

Full inclusion and exclusion criteria are listed as follows:

*Inclusion Criteria*

1. Written informed consent
2. Healthy men or women, aged ≥ 18 ≤ 60 at day of inclusion having received either
   a) primary immunization (cohort 1)† with any regimen using any EU marketed SARS-CoV-2 vaccine or
   b) subsequently booster immunization (cohort 2) with any EU marketed mRNA vaccine at least 3 months prior to enrolment
3. Adults with SARS-CoV-2 specific IgG concentration between 10 RU/mL and 1200 RU/mL determined by Anti-SARS-CoV-2-QuantiVac-ELISA (IgG)
4. Males or non-pregnant, non-lactating females of child-bearing potential with negative pregnancy test at screening who agree to comply with the applicable contraceptive requirements of the protocol from at least 14 days prior to vaccination and during the entire duration of the study or females without child-bearing potential
5. Normal pulmonary function: FEV1 predicted ≥ 80% and FEV1/FVC > 70%
6. Body mass index 18.5 - 30.0 kg/ m2 and weight > 50 kg at screening

7. Participant is capable of understanding the investigational nature, potential risks and benefits of the clinical trial

*Exclusion Criteria*

1. Previous MVA or rMVA vaccination
2. Known allergy to the components of the SARS-CoV-2 vaccine product such as chicken proteins or history of life-threatening reactions to vaccine containing the same substances
3. Known history of anaphylaxis to vaccination or any allergy likely to be exacerbated by any component of the trial vaccine
4. Any laboratory value outside the reference range that the investigator considers to be of clinical relevance; safety laboratory screening evaluation can be repeated a maximum of two times
5. Any finding in the medical history and physical examination deviating from normal and assessed as clinically relevant by the investigator
6. Evidence in the participant's medical history or in the medical examination that might influence the absorption, distribution, metabolism or excretion of the investigational medicinal product
7. Current smoking/ vaping or smoking /vaping in the previous year.
8. Clinically relevant findings in ECG
9. Any confirmed or suspected immunosuppressive or immunodeficient condition, cytotoxic therapy in the previous 5 years, and/or diabetes
10. Asthma, chronic obstructive pulmonary disease or other lung disease
11. Respiratory tract infection in the 4 weeks prior to study treatment
12. Any chronic or active neurologic disorder, including seizures, and epilepsy, excluding a single febrile seizure as a child
13. Known intolerance to medication used during bronchoscopy, i.e. midazolam and lidocaine.
14. Treatment with ß-adrenoceptor antagonists

15. Alcohol abuse (consumption of more than 20 g per day for females and 30 g per day for males)
16. Drug abuse or positive drug screening
17. Any positive result for HIV1/2, HCV antibody or HBs antigen testing
18. Moderate or severe illness and/or fever >38 °C within 1 week prior to vaccination
19. History of blood donation within 60 days of enrollment or plans to donate within the treatment phase
20. Participation in a clinical trial or use of an investigational product within 30 days or five times the half-life of the investigational product -whichever is longer- prior to receiving the first dose within this study
21. Investigator or employee of the study site or Sponsor with direct involvement in the proposed study, or identified as an immediate family member (i.e. parent, natural or adopted child) of the investigator or employee with direct involvement in the proposed study

*† Cohort 1 was planned to enroll adults with primary but not booster SARS-CoV-2 immunization. However, eligible participants could not be identified due to rapid and widespread administration of booster doses during the evolving pandemic. The study proceeded exclusively with Cohort 2; no participants were enrolled in Cohort 1.*

#### Supplementary Information S2: Aerosol Stability Assessment of Study Vaccine

Given that MVA-SARS-2-S and MVA-SARS-2-ST share an identical poxviral backbone, in-use aerosol stability studies were carried out with MVA-SARS-2-S, serving as a surrogate for the clinical vaccine candidate. To assess the integrity of the vaccine following aerosolization, five vials (each containing 0.5 mL of 1x $10^7$ IU) were aerosolized into a 50-mL collection tube pre-filled with 19.5 mL growth medium. The nebulizer unit was operated without a mouthpiece, and the remaining virus in the vial was kept on ice to serve as a non-nebulized infectivity control. The aerosolization proceeded in cycles: 10 seconds of nebulization, gentle swinging of the collection tube for 5 seconds to ensure uniform distribution, and 20 seconds on ice. This cycle was repeated until the entire volume was nebulized.

Infectivity of both nebulized and non-nebulized samples was assessed by titration on avian DF-1 cells. Specifically, $1x10^6$ DF-1 cells were seeded per well in 6-well plates and incubated for 4 hours (±15 minutes) at 37°C with 5% $CO_2$. Following incubation, the cells were inoculated with 0.5 mL of diluted virus preparation or, for negative controls, with medium alone. Viral infection was allowed to proceed for 15 hours (±60 minutes) at 37°C and 5% $CO_2$. After the infection period, cells were harvested by brief treatment with TrypLE Select (Gibco), then fixed and permeabilized using BD Cytofix/Cytoperm reagent. Intracellular viral protein detection was performed by staining with a FITC-conjugated anti-vaccinia antibody (BioSynth, #60-V68). Samples were analyzed by flow cytometry using a BD LSR II Flow Cytometer, with only those virus dilutions resulting in 6.0-53.0% infected cells included in subsequent titer calculations, using the following formula:

$$\text{Infectious titer } \left[\frac{\text{IU}}{\text{mL}}\right] = \frac{\sum\left(\left(\frac{\text{infected cells [\%]}}{100}\right) \text{x } \left(\frac{\text{DF}}{\text{K}}\right) \text{x D}\right)}{\text{n}}$$

where DF is the dilution factor, K is the inoculation volume (0.5 mL), D is the seed density (1 x 10^6 cells/well), and n is the number of evaluable dilutions.

#### Supplementary Table S1: MVA-SARS-2-ST Vaccine Titers and Delivered Dose Following Nebulization

| | **Volume** | **MVA-SARS-2-ST titer** |
|---|---|---|
| Drug product filled titer | 1 mL | $3.34 \times 10^{7}$ IU/mL |
| Nominal titer per dose | 0.5 mL | $1.67 \times 10^{7}$ IU/dose |
| Effective dose (after nebulization of 0.5 mL IMP) | 0.5 mL | $1.00 \times 10^{7}$ IU/dose |

#### Supplementary Table S2: Listing of all AEs throughout study period, sorted by participant

| Participant ID* | MedDRA preferred term | Solicited adverse event? | Severity | Grading for local AEs, physical observations and systemic AEs | Causality assessment | AE onset (days since inhalation boost) | Duration of AE | Outcome |
|---|---|---|---|---|---|---|---|---|
| P001 | Dermatitis allergic | no | moderate | Grade 2 | Unrelated | 39 | 28 | Recovered / Resolved |
| P001 | Palpitations | no | mild | Grade 1 | Possible | 0 | 1 | Recovered / Resolved |
| P002 | Nasopharyngitis | no | mild | Grade 1 | Unrelated | 108 | 8 | Recovered / Resolved |
| P002 | Cough | yes | mild | Grade 1 | Very likely / certain | 0 | 7 | Recovered / Resolved |
| P002 | Hypertension | no | mild | Grade 1 | Unlikely | 0 | 1 | Recovered / Resolved |
| P003 | Hypertension | no | mild | Grade 1 | Unrelated | 144 | . | Ongoing |
| P003 | Dark circles under eyes | no | mild | Grade 1 | Unlikely | 9 | 1 | Recovered / Resolved |
| P003 | Fatigue | no | moderate | Grade 2 | Unrelated | 8 | 1 | Recovered / Resolved |
| P003 | Conjunctival haemorrhage | no | mild | Grade 1 | Unlikely | 32 | 2 | Recovered / Resolved |
| P003 | Conjunctival haemorrhage | no | mild | Grade 1 | Unlikely | 37 | 3 | Recovered / Resolved |
| P003 | Lacrimation increased | no | mild | Grade 1 | Unlikely | 9 | 1 | Recovered / Resolved |
| P003 | Photophobia | no | mild | Grade 1 | Unlikely | 9 | 1 | Recovered / Resolved |
| P003 | Decreased appetite | no | mild | Grade 1 | Possible | 1 | 1 | Recovered / Resolved |
| P003 | Sensation of foreign body | no | mild | Grade 1 | Possible | 1 | 1 | Recovered / Resolved |
| P003 | Cough | no | mild | Grade 1 | Unrelated | 12 | 1 | Recovered / Resolved |

| Participant ID* | MedDRA preferred term | Solicited adverse event? | Severity | Grading for local AEs, physical observations and systemic AEs | Causality assessment | AE onset (days since inhalation boost) | Duration of AE | Outcome |
|---|---|---|---|---|---|---|---|---|
| P003 | Fatigue | no | mild | Grade 1 | Unrelated | 12 | 1 | Recovered / Resolved |
| P003 | Procedural headache | no | mild | Grade 1 | Unrelated | 12 | 1 | Recovered / Resolved |
| P003 | Muscle spasms | no | severe | Grade 3 | Unrelated | 54 | 1 | Recovered / Resolved |
| P003 | Nasal congestion | no | mild | Grade 1 | Unrelated | 12 | 10 | Recovered / Resolved |
| P003 | Procedural pain | no | moderate | Grade 2 | Very likely / certain | 11 | 66 | Recovered / Resolved |
| P003 | Respiratory tract infection | no | moderate | Grade 1 | Unlikely | 115 | 13 | Recovered / Resolved |
| P003 | Hypertension | no | mild | Grade 1 | Unlikely | 11 | .* | Recovered / Resolved |
| P003 | Dyspnoea | no | mild | Grade 1 | Unrelated | 8 | 2 | Recovered / Resolved |
| P003 | Post procedural fever | no | mild | Grade 1 | Unrelated | 12 | 1 | Recovered / Resolved |
| P003 | Rhonchi | no | mild | Grade 1 | Unclassifiable | 77 | 5 | Recovered / Resolved |
| P003 | Oropharyngeal pain | no | mild | Grade 1 | Unrelated | 92 | 3 | Recovered / Resolved |
| P004 | Chills | no | moderate | Grade 2 | Unrelated | 14 | 1 | Recovered / Resolved |
| P004 | COVID-19 | no | mild | Grade 1 | Unrelated | 97 | 6 | Recovered / Resolved |
| P004 | Cough | yes | moderate | Grade 2 | Possible | 5 | 2 | Recovered / Resolved |
| P004 | Hypertension | no | mild | Grade 1 | Unrelated | 137 | . | Ongoing |

| Participant ID* | MedDRA preferred term | Solicited adverse event? | Severity | Grading for local AEs, physical observations and systemic AEs | Causality assessment | AE onset (days since inhalation boost) | Duration of AE | Outcome |
|---|---|---|---|---|---|---|---|---|
| P004 | Fatigue | yes | mild | Grade 1 | Probable | 5 | 2 | Recovered / Resolved |
| P004 | Chest discomfort | yes | moderate | Grade 2 | Probable | 4 | 2 | Recovered / Resolved |
| P004 | Pyrexia | yes | moderate | Grade 2 | Possible | 5 | 2 | Recovered / Resolved |
| P004 | Post procedural fever | no | moderate | Grade 2 | Unrelated | 14 | 1 | Recovered / Resolved |
| P004 | Headache | yes | mild | Grade 1 | Probable | 5 | 2 | Recovered / Resolved |
| P004 | Headache | no | mild | Grade 1 | Unrelated | 10 | 2 | Recovered / Resolved |
| P004 | Influenza | no | mild | Grade 1 | Unlikely | 98 | 8 | Recovered / Resolved |
| P005 | Cough | no | mild | Grade 1 | Unlikely | 14 | 4 | Recovered / Resolved |
| P005 | Fatigue | yes | mild | Grade 1 | Possible | 0 | 2 | Recovered / Resolved |
| P005 | Fatigue | no | mild | Grade 1 | Unlikely | 14 | 2 | Recovered / Resolved |
| P005 | Headache | no | moderate | Grade 2 | Unlikely | 15 | 2 | Recovered / Resolved |
| P005 | Myalgia | yes | mild | Grade 1 | Possible | 2 | 3 | Recovered / Resolved |
| P005 | Myalgia | no | moderate | Grade 2 | Unrelated | 14 | . | Not recovered / not resolved |
| P005 | Myalgia | no | mild | Grade 1 | Unlikely | 14 | 2 | Recovered / Resolved |
| P006 | COVID-19 | no | moderate | Grade 2 | Unrelated | 65 | 15 | Recovered / Resolved |
| P006 | Brain fog | no | mild | Grade 1 | Possible | 0 | 1 | Recovered / Resolved |
| P006 | Colonoscopy | no | moderate | Grade 2 | Unrelated | 109 | 1 | Recovered / Resolved |

| Participant ID* | MedDRA preferred term | Solicited adverse event? | Severity | Grading for local AEs, physical observations and systemic AEs | Causality assessment | AE onset (days since inhalation boost) | Duration of AE | Outcome |
|---|---|---|---|---|---|---|---|---|
| P007 | Back pain | no | moderate | Grade 2 | Unrelated | 126 | 8 | Recovered / Resolved |
| P007 | Fatigue | no | moderate | Grade 2 | Unrelated | 22 | 3 | Recovered / Resolved |
| P007 | Oral herpes | no | moderate | Grade 2 | Unrelated | 134 | 15 | Recovered / Resolved |
| P007 | Headache | no | moderate | Grade 2 | Possible | 21 | 4 | Recovered / Resolved |
| P007 | Nasopharyngitis | no | moderate | Grade 2 | Unrelated | 93 | 2 | Recovered / Resolved |
| P007 | Sensitive skin | no | mild | Grade 1 | Unlikely | 21 | 23 | Recovered / Resolved |
| P008 | COVID-19 | no | moderate | Grade 2 | Unrelated | 99 | 11 | Recovered / Resolved |
| P008 | Cough | yes | mild | Grade 1 | Possible | 1 | 16 | Recovered / Resolved |
| P008 | Nasopharyngitis | no | mild | Grade 1 | Unrelated | 67 | 11 | Recovered / Resolved |
| P009 | Nasopharyngitis | no | moderate | Grade 2 | Unlikely | 12 | 11 | Recovered / Resolved |
| P009 | Urinary tract infection | no | mild | Grade 1 | Unrelated | 143 | . | Not recovered / not resolved |
| P010 | Pulmonary function test decreased | no | mild | Grade 1 | Possible | 14 | 29 | Recovered / Resolved |
| P010 | Procedural dizziness | no | mild | Grade 1 | Unrelated | 14 | 1 | Recovered / Resolved |
| P010 | Throat irritation | no | mild | Grade 1 | Unrelated | 14 | 4 | Recovered / Resolved |
| P010 | Cough | no | mild | Grade 1 | Unrelated | 28 | 6 | Recovered / Resolved |
| P011 | Nasopharyngitis | no | mild | Grade 1 | Unrelated | 53 | 4 | Recovered / Resolved |

| Participant ID* | MedDRA preferred term | Solicited adverse event? | Severity | Grading for local AEs, physical observations and systemic AEs | Causality assessment | AE onset (days since inhalation boost) | Duration of AE | Outcome |
|---|---|---|---|---|---|---|---|---|
| P011 | Artificial crown procedure | no | moderate | Grade 2 | Unrelated | 84 | 1 | Recovered / Resolved |
| P011 | Pain in extremity | no | moderate | Grade 2 | Unrelated | 91 | 10 | Recovered / Resolved |
| P011 | Headache | no | moderate | Grade 2 | Unrelated | 94 | 7 | Recovered / Resolved |
| P011 | Cough | no | mild | Grade 1 | Possible | .[#] | . | Not recovered / not resolved |
| P011 | Nasal congestion | no | mild | Grade 1 | Possible | 3 | 42 | Recovered / Resolved |
| P011 | Vascular pain | no | mild | Grade 1 | Probable | .[#] | . | Not recovered / not resolved |
| P011 | Pyrexia | yes | mild | Grade 1 | Possible | 2 | 3 | Recovered / Resolved |
| P011 | Abdominal pain upper | yes | mild | Grade 1 | Unlikely | 1 | 1 | Recovered / Resolved |
| P011 | Abdominal pain upper | no | mild | Grade 1 | Unrelated | 16 | 1 | Recovered / Resolved |
| P012 | Muscle tightness | no | mild | Grade 1 | Unrelated | 15 | 2 | Recovered / Resolved |
| P013 | Cough | no | moderate | Grade 2 | Unrelated | 18 | 5 | Recovered / Resolved |
| P013 | Cough | no | mild | Grade 1 | Unrelated | 23 | 14 | Recovered / Resolved |
| P013 | Gastrointestinal infection | no | moderate | Grade 2 | Unrelated | 83 | 19 | Recovered / Resolved |
| P013 | Arthralgia | no | mild | Grade 1 | Unrelated | 55 | 1 | Recovered / Resolved |
| P013 | Productive cough | no | mild | Grade 1 | Unrelated | 16 | 1 | Recovered / Resolved |
| P013 | Bradycardia | no | mild | Grade 1 | Unrelated | 0 | 1 | Recovered / Resolved |

| Participant ID* | MedDRA preferred term | Solicited adverse event? | Severity | Grading for local AEs, physical observations and systemic AEs | Causality assessment | AE onset (days since inhalation boost) | Duration of AE | Outcome |
|---|---|---|---|---|---|---|---|---|
| P014 | Fatigue | yes | mild | Grade 1 | Unlikely | 7 | 3 | Recovered / Resolved |
| P014 | Headache | yes | mild | Grade 1 | Unlikely | 7 | 3 | Recovered / Resolved |
| P014 | Bundle branch block right | no | mild | Grade 1 | Unrelated | 150 | . | Not recovered / not resolved |
| P014 | Upper respiratory tract infection | no | mild | Grade 1 | Unrelated | 124 | 31 | Recovered / Resolved |
| P014 | Throat irritation | yes | mild | Grade 1 | Very likely / certain | 0 | 1 | Recovered / Resolved |
| P014 | Hypertension | no | mild | Grade 1 | Unrelated | 1 | 3 | Recovered / Resolved |
| P014 | Back pain | no | mild | Grade 1 | Unrelated | 26 | 4 | Recovered / Resolved |
| P014 | Arthropod bite | no | mild | Grade 1 | Unrelated | 12 | 3 | Recovered / Resolved |
| P015 | Cough | no | mild | Grade 1 | Possible | 14 | 22 | Recovered / Resolved |
| P015 | Hypertension | no | mild | Grade 1 | Unlikely | 3 | 12 | Recovered / Resolved |
| P015 | Pyrexia | no | mild | Grade 1 | Possible | 14 | 2 | Recovered / Resolved |
| P015 | Headache | yes | moderate | Grade 2 | Possible | 3 | 1 | Recovered / Resolved |
| P015 | Malaise | yes | moderate | Grade 2 | Possible | 3 | 1 | Recovered / Resolved |
| P015 | Flushing | no | mild | Grade 1 | Possible | 3 | 1 | Recovered / Resolved |
| P015 | Flushing | no | mild | Grade 1 | Possible | 0 | 1 | Recovered / Resolved |
| P015 | Headache | yes | mild | Grade 1 | Possible | 0 | 2 | Recovered / Resolved |
| P015 | Chest pain | yes | mild | Grade 1 | Possible | 0 | 1 | Recovered / Resolved |

| Participant ID* | MedDRA preferred term | Solicited adverse event? | Severity | Grading for local AEs, physical observations and systemic AEs | Causality assessment | AE onset (days since inhalation boost) | Duration of AE | Outcome |
|---|---|---|---|---|---|---|---|---|
| P015 | Pyrexia | yes | mild | Grade 1 | Possible | 3 | 1 | Recovered / Resolved |
| P015 | Pyrexia | yes | mild | Grade 1 | Possible | 0 | 1 | Recovered / Resolved |
| P015 | Diarrhoea infectious | no | mild | Grade 1 | Unrelated | 115 | 5 | Recovered / Resolved |
| P016 | Abdominal pain | yes | mild | Grade 1 | Possible | 1 | 3 | Recovered / Resolved |
| P016 | Nasopharyngitis | no | moderate | Grade 2 | Unrelated | 54 | 13 | Recovered / Resolved |
| P016 | Dizziness | no | moderate | Grade 2 | Unlikely | 14 | 1 | Recovered / Resolved |
| P016 | Cough | yes | mild | Grade 1 | Probable | 1 | 1 | Recovered / Resolved |
| P016 | Cough | no | moderate | Grade 2 | Probable | 77 | 11 | Recovered / Resolved |
| P016 | Fatigue | yes | mild | Grade 1 | Probable | 0 | 1 | Recovered / Resolved |
| P016 | Headache | yes | mild | Grade 1 | Probable | 0 | 1 | Recovered / Resolved |
| P016 | Headache | no | mild | Grade 1 | Unlikely | 14 | 2 | Recovered / Resolved |
| P016 | Cough | yes | mild | Grade 1 | Probable | 0 | 1 | Recovered / Resolved |
| P016 | Nasal congestion | no | mild | Grade 1 | Possible | 1 | 3 | Recovered / Resolved |
| P016 | Oropharyngeal pain | no | mild | Grade 1 | Unrelated | 28 | 3 | Recovered / Resolved |
| P017 | Epistaxis | no | mild | Grade 1 | Unlikely | 2 | 1 | Recovered / Resolved |
| P017 | Headache | yes | mild | Grade 1 | Possible | 3 | 1 | Recovered / Resolved |

| Participant ID* | MedDRA preferred term | Solicited adverse event? | Severity | Grading for local AEs, physical observations and systemic AEs | Causality assessment | AE onset (days since inhalation boost) | Duration of AE | Outcome |
|---|---|---|---|---|---|---|---|---|
| P017 | Alcoholic hangover | no | moderate | Grade 2 | Possible | 97 | 1 | Recovered / Resolved |
| P017 | Headache | yes | mild | Grade 1 | Probable | 0 | 1 | Recovered / Resolved |
| P017 | Ear pain | no | mild | Grade 1 | Unlikely | 11 | 1 | Recovered / Resolved |
| P017 | Hyperhidrosis | no | mild | Grade 1 | Possible | 3 | 1 | Recovered / Resolved |
| P018 | Nasopharyngitis | no | moderate | Grade 1 | Unlikely | 32 | 9 | Recovered / Resolved |
| P018 | Nasopharyngitis | no | mild | Grade 1 | Unrelated | 120 | 9 | Recovered / Resolved |
| P018 | Rhinitis | no | mild | Grade 1 | Unrelated | 92 | 5 | Recovered / Resolved |
| P018 | Dry throat | yes | mild | Grade 1 | Unlikely | 0 | 1 | Recovered / Resolved |
| P018 | Rhinorrhoea | no | mild | Grade 1 | Unlikely | 1 | 1 | Recovered / Resolved |
| P018 | Muscle strain | no | moderate | Grade 2 | Unrelated | 35 | 8 | Recovered / Resolved |
| P019 | Nasopharyngitis | no | mild | Grade 1 | Unclassifiable | 19 | 5 | Recovered / Resolved |
| P019 | Fatigue | yes | mild | Grade 1 | Possible | 1 | 1 | Recovered / Resolved |
| P019 | Pyrexia | no | mild | Grade 1 | Unlikely | 14 | 2 | Recovered / Resolved |
| P019 | Cough | no | mild | Grade 1 | Unrelated | 14 | 6 | Recovered / Resolved |
| P019 | Throat irritation | yes | mild | Grade 1 | Possible | 1 | 2 | Recovered / Resolved |
| P019 | Electrocardiogram QRS complex shortened | no | mild | Grade 1 | Unrelated | 137 | . | Not recovered / not resolved |

| Participant ID* | MedDRA preferred term | Solicited adverse event? | Severity | Grading for local AEs, physical observations and systemic AEs | Causality assessment | AE onset (days since inhalation boost) | Duration of AE | Outcome |
|---|---|---|---|---|---|---|---|---|
| P019 | Cough | yes | mild | Grade 1 | Possible | 0 | 1 | Recovered / Resolved |
| P019 | Pyrexia | yes | mild | Grade 1 | Possible | 1 | 1 | Recovered / Resolved |
| P020 | Headache | no | mild | Grade 1 | Unrelated | 14 | 1 | Recovered / Resolved |
| P020 | Headache | no | moderate | Grade 2 | Unrelated | 74 | 3 | Recovered / Resolved |
| P020 | Headache | no | moderate | Grade 2 | Unrelated | 93 | 3 | Recovered / Resolved |
| P020 | Procedural headache | no | mild | Grade 1 | Unrelated | 8 | 2 | Recovered / Resolved |
| P020 | Tachycardia paroxysmal | no | moderate | Grade 2 | Unlikely | 62 | 4 | Recovered / Resolved |
| P020 | Restless legs syndrome | no | moderate | Grade 2 | Unrelated | 14 | 1 | Recovered / Resolved |
| P020 | Sinusitis | no | moderate | Grade 2 | Unrelated | 133 | 12 | Recovered / Resolved |
| P020 | Fatigue | yes | mild | Grade 1 | Probable | 0 | 1 | Recovered / Resolved |
| P020 | Hyperhidrosis | yes | mild | Grade 1 | Possible | 1 | 1 | Recovered / Resolved |
| P020 | Hyperhidrosis | no | mild | Grade 1 | Unlikely | 30 | 6 | Recovered / Resolved |
| P020 | Hyperhidrosis | no | mild | Grade 1 | Unlikely | 62 | 4 | Recovered / Resolved |
| P020 | Tachycardia | no | moderate | Grade 2 | Unlikely | 30 | 6 | Recovered / Resolved |
| P020 | Fatigue | no | mild | Grade 1 | Unrelated | 8 | 1 | Recovered / Resolved |

| **Participant ID*** | **MedDRA preferred term** | **Solicited adverse event?** | **Severity** | **Grading for local AEs, physical observations and systemic AEs** | **Causality assessment** | **AE onset (days since inhalation boost)** | **Duration of AE** | **Outcome** |
|---|---|---|---|---|---|---|---|---|
| P021 | Nasopharyngitis | no | mild | Grade 1 | Unrelated | 99 | 5 | Recovered / Resolved |
| P021 | Epistaxis | no | severe | Grade 2 | Unlikely | 28 | 1 | Recovered / Resolved |
| P021 | Myalgia | no | mild | Grade 1 | Unrelated | 133 | . | Not recovered / not resolved |
| P021 | Urinary tract infection | no | moderate | Grade 2 | Unrelated | 27 | 2 | Recovered / Resolved |

* Identifiers were regenerated for publication and are pseudonymized codes only. Mapping to original study IDs is not included in the published materials.

### AE start date partially missing (day of date unknown). According to the available information on month and year of AE start date, the AE onset was between 61 and 90 days after inhalation boost

#### Supplementary Table S3: Levels of S1-binding IgG antibodies in blood - Absolute change (difference) from baseline

| | n | Mean [95% CI] | SD | CV | [Min, Max] | Median | IQR |
|---|---|---|---|---|---|---|---|
| **Difference in S1-binding IgG in blood (BAU/mL) from baseline** | | | | | | | |
| to day 7 | 23 | 34.7 [-83.1, 152.4] | 272.3 | 7.9 | [-477, 747] | -2.0 | 180.0 |
| to day 14 | 22 | 16.0 [-177.4, 209.4] | 436.1 | 27.3 | [-885, 1315] | -30.0 | 200.0 |
| to day 28 | 23 | 130.8 [-138.3, 399.9] | 622.3 | 4.8 | [-504, 2562] | -1.0 | 178.0 |
| to day 56 | 23 | 25.6 [-150.5, 201.6] | 407.1 | 15.9 | [-1134, 862] | 27.0 | 396.0 |
| to day 140 | 23 | 316.7 [-409.5, 1042.8] | 1679.1 | 5.3 | [-777, 7350] | -62.0 | 535.0 |

n: Number of nonmissing values; CI: Confidence interval; SD: Standard Deviation; CV: Coefficient of Variation; Min: Minimum; Max: Maximum; IQR: Interquartile Range

#### Supplementary Table S4: Levels of S1-binding IgG antibodies in bronchoalveolar lavage (BAL) - Absolute change (difference) from baseline

| | n | Mean [95% CI] | SD | CV | [Min, Max] | Median | IQR |
|---|---|---|---|---|---|---|---|
| **Difference in S1-binding IgG in BAL (BAU) from baseline** | | | | | | | |
| to day 14 | 22 | 6.0 [-22.8, 34.7] | 64.7 | 10.9 | [-182, 200] | 4.5 | 21.0 |

n: Number of nonmissing values; CI: Confidence interval; SD: Standard Deviation; CV: Coefficient of Variation; Min: Minimum; Max: Maximum; IQR: Interquartile Range

#### Supplementary Table S5: Levels of S1-binding IgA antibodies in blood - Absolute change (difference) from baseline

| | n | Mean [95% CI] | SD | CV | [Min, Max] | Median | IQR |
|---|---|---|---|---|---|---|---|
| **Difference in S1-binding IgA in blood (MFI) from baseline** | | | | | | | |
| to day 7 | 23 | 20.6 [-3.7, 44.9] | 56.2 | 2.7 | [-55, 171] | 1.0 | 43.0 |
| to day 14 | 22 | 34.4 [-8.8, 77.5] | 97.3 | 2.8 | [-166, 295] | 14.0 | 78.4 |
| to day 28 | 23 | 33.5 [3.7, 63.4] | 69.1 | 2.1 | [-58, 193] | 20.0 | 63.0 |
| to day 56 | 23 | 30.7 [-19.6, 80.9] | 116.2 | 3.8 | [-270, 322] | 1.0 | 57.5 |
| to day 140 | 23 | 43.1 [-67.0, 153.2] | 254.6 | 5.9 | [-320, 782] | -4.0 | 176.0 |

n: Number of nonmissing values; CI: Confidence interval; SD: Standard Deviation; CV: Coefficient of Variation; Min: Minimum; Max: Maximum; IQR: Interquartile Range

#### Supplementary Table S6: Levels of S1-binding IgA antibodies in bronchoalveolar lavage (BAL) - Absolute change (difference) from baseline

| | n | Mean [95% CI] | SD | CV | [Min, Max] | Median | IQR |
|---|---|---|---|---|---|---|---|
| **Difference in S1-binding IgA in BAL (MFI) from baseline** | | | | | | | |
| to day 14 | 22 | 40.5 [7.5, 73.5] | 74.5 | 1.8 | [-128, 215] | 5.3 | 68.8 |

n: Number of nonmissing values; CI: Confidence interval; SD: Standard Deviation; CV: Coefficient of Variation; Min: Minimum; Max: Maximum; IQR: Interquartile Range

#### Supplementary Table S7: Study participants with reported or suspected COVID-19 breakthrough infections

| Participant ID* | Timepoint of reporting COVID-19 as adverse event | Positive PCR test for SARS-CoV-2 | S1-IgG titer development after (suspected) breakthrough infection | NCP-IgG titer status after (suspected) breakthrough infection |
|---|---|---|---|---|
| P004 | Day 97 | No | ↑ increase (rise from 392 to 2551 BAU/mL) | seronegative |
| P006 | Day 65 | Yes (Day 140) | ↑increase (rise from 155 to 7544 BAU/mL) | seropositive |
| P008 | Day 99 | No | → no relevant change (change from 774 to 701 BAU/mL) | seropositive |
| P022 | No COVID-19 reported | No | ↑increase (rise from 2198 to 3422 BAU/mL) | seropositive |

* Identifiers were regenerated for publication and are pseudonymized codes only. Mapping to original study IDs is not included in the published materials.

#### Supplementary Figure S1: Serum NCP-specific IgG from day 0 to day 140 following inhaled MVA-SARS-2-ST

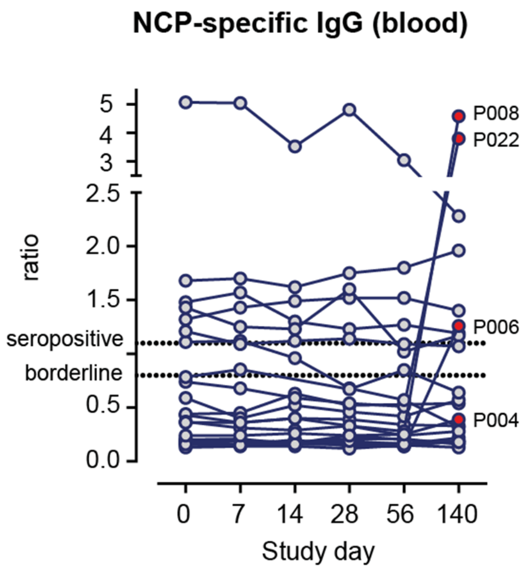


Antibody levels were determined with a semi-quantitative Anti-SARS-CoV-2-NCP-IgG ELISA. The NCP ratio corresponds to the absorbance of the sample divided through the absorbance of the reference calibrator. Ratio < 0.8: seronegative; Ratio ≥ 0.8 to < 1.1: borderline; Ratio ≥ 1.1: seropositive. Red-filled circles labeled with IDs indicate reported or suspected COVID-19 breakthrough infection.
